# Is the health system ready for future crises? A qualitative study protocol

**DOI:** 10.1101/2022.11.27.22282798

**Authors:** Hania Rahimi-Ardabili, Farah Magrabi, Enrico Coiera

**Affiliations:** Centre for Health Informatics, Australian Institute of Health Innovation, Macquarie University, Australia

## Abstract

The COVID-19 pandemic serves as a clarion call to ensure health systems are better prepared to meet future emergencies. Digital Health could play a significant role in preparing health systems to bend and stretch their resources and cope with various shocks by facilitating tasks such as disease monitoring and care delivery. However, the health system’s needs during the crises have not been thoroughly examined from the perspective of health professionals in general and in the Australian health setting in particular. Here we describe the protocol of a qualitative design to learn from frontline healthcare workers’ experiences of the pandemic response that can guide preparation for future crises using digital health.

## Introduction

COVID-19 poses a great burden on public health systems worldwide as many were overwhelmed by a quickly spreading new coronavirus (Sheehan and Fox 2020). This pandemic serves as a clarion call to ensure health systems are better prepared to meet the emergencies of the future. The health system needs to be engineered in a way that bends and stretches its resources and capabilities quickly and repeatedly to cope with various shocks from climate change-related crises (Coiera and Braithwaite 2021). We might call this a ‘turbulence system’—one purpose-designed to operate across a wide variety of conditions and to efficiently and effectively reconfigure itself to meet different demands. Health Informatics has already assisted the health system with monitoring and managing the care delivery process (Kelly, Campbell et al. 2020, Sarbadhikari and Sarbadhikari 2020), and managing disease outbreaks (Golinelli, Boetto et al. 2020) could play a significant role in achieving these aims. Artificial intelligence and digital services can allow us to collect large amounts of data from various resources, such as environmental data from sensors and remote sensing satellites, and analyse using powerful analytical models to transform them into understandable data for decision-making and tracking environmental changes to project future. A work from Campbell et al. is an example of a use of a mobile app that analysis information to detect air quality, tracks user symptoms in near real-time, and assists vulnerable populations in reducing their smoke exposure and protecting their health (Campbell, Jones et al. 2020). However, how digital services can assist health systems and health professionals and what tools and strategies health systems should adopt to be turbulence-ready is not thoroughly examined from the perspective of health professionals in general and in the Australian health setting in particular. Learning from the experience of the pandemic response can provide some guidance on preparing for future crises and how health informatics could be a solution for it from frontline healthcare workers’ perspective.

## Methods

### Design

We propose a qualitative approach to explore frontline healthcare workers’ lived experiences with the potential challenges in the health system. Qualitative research is the most suitable approach when there is little knowledge about the phenomenon. A one-on-one semi-structured interview will be utilised for this purpose. It provides in-depth information about frontline healthcare workers’ needs and their perspective on how informatics technology can help build a turbulence-ready health system.

### Setting

Participants will be health professionals working in various settings in the health system, including hospitals, general practice, public health, laboratory, NSW health, government, and private settings.

### Sample size and recruitment

Recruitment will continue until rich information is attained. We anticipate we will achieve thematic saturation by recruiting approximately 25 participants. A systematic review of qualitative studies conducted about healthcare frontline workers’ experience with pandemics reported that 14 studies that conducted one-on-one interviews recruited, on average, 15 participants each (Billings, Ching et al. 2021).

A purposive and convenient sampling approach will be taken to capture a wide range of experiences from different roles and levels. For this purpose, the research team (E.C., B.S., T.S.), will identify eligible individuals using their Networks. The snowballing method will also be performed. Participants may be asked to email the study invitation email and participant information to their colleagues and networks who may be interested.

### Ethics and consent

Ethical approval has been sought before recruitment and data collection from the Macquarie University Ethics Committee. Participants will be asked to indicate their written consent before any data collection.

#### Data collection procedures

Interviews will be conducted online using the Zoom videoconference platform. Interviews will be conducted by one of the research team (H.R-A.) experienced in qualitative interviews. Each interview will take about 30-40 minutes. An automated transcript function will be used, and the research team will perform a random cross-check of transcripts for accuracy. Data collection and analysis will occur concurrently, and emerging themes from initial data analysis may shape subsequent interview questions and samplings.

### Participants

Participants will be clinicians involved in the pandemic response, including frontline healthcare workers (from public and private sectors) such as:

- emergency department clinicians, including intensive care unit (ICU) specialists;
- general practitioners (from various settings, hospitals, public health, primary care, and aged care);
- nurses (hospital nurse, community nurse, nurse manager),
- pathologists (infectious disease specialists);
- allied health care (e.g. physiologists working in ICUs);
- management decision-makers, NSW health officer, executive teams in government;
- administrative staff in hospitals;
- midwives;
- hospital medical officers,
- other aged care workers (facility managers, registered nurses, assistants in nursing (AINs) roles);
- respiratory specialists;
- paramedics.

### Data analysis

transcripts will be qualitatively analysed using reflexive thematic analysis to determine key themes identified by participants (Clarke, Braun et al. 2015, Braun and Clarke 2019). A couple of initial transcripts will be open-coded line-by-line coding. To ensure credibility, researcher triangulation will be performed, meaning that a few initial transcripts will be independently coded by research teams, and codes will be then discussed. One investigator (H.R-A.) will analyse the rest of the data using both inductive and deductive approaches. Data analysis will be performed using NVivo Software. Codebooks will be developed and will be refined (some codes collapsed or separated) as higher-level themes emerge. Modified emerging codes and themes will be discussed further with all investigators for credibility. The main analyst (H.R-A.) will develop memos for analysis and an audit trail.

Participants will be offered an opportunity to review the initial themes to check if they reflect their voices if they wish. Their feedback will further be incorporated into the analysis. Results will be presented using various de-identified quotations from participants to reflect their voices. A novel framework will be developed and will be supported by multiple theories (Theoretical triangulation).

### Reflexivity

Authors (E.C., B.S., T.S.) have a clinical background with two in current clinical practice (B.S., T.S.), and all authors are experienced and involved in research activities relevant to the health system. All authors had prior experience in analysing qualitative data. The interviewer and main analyst (H.R-A.) has not previously practised in Australian health service and do not have any prior contact or relationship with any of the participants thus; the analyst’s lack of experience balances questioning of positive and negative experiences to explore and interpret data without bias or assumptions regarding the data. In addition, she will keep a reflexivity journal during analysis to discuss it with all authors.

### Assuring rigour

A number of approaches will be taken to ensure credibility, transferability, dependability and confirmability. The techniques used to ensure rigour are summarised in the table.

**Table.** Strategies to ensure rigour

| Rigour criteria | Strategies to ensure criteria are met |
| --- | --- |
| Credibility | Purposeful sampling<br>Member checking by providing an opportunity for participants to provide feedback on initial themes<br>Investigator triangulation by involving more than one analyst<br>Data triangulation by recruiting participants from various health professions and settings<br>Theoretical triangulation by bringing more than one theory for supporting findings<br>Audit trail of coding decisions and data analysis procedures using codebook and memos<br>Quotations from participants to support the interpretation |
| Transferability | Purposeful sampling<br>Thick description of research setting, and methods used in the research |
| Dependability | Member checking<br>Investigator, data and theoretical triangulation<br>Audit trail |
| Confirmability | Audit trail of coding decisions and data analysis procedures using codebook and memos |

## Discussion

### Strengths and limitations

While we aim to recruit a wide range of health professionals from different settings using purpose sampling, however; considering that the recruitment will be done through using the investigators’ network, there is a chance that participants share a similar opinion. Similarly, snowballing could result in recruiting participants with a similar perspective as they are from same networks.

### Expected output

We expect data coming from this study will aid in developing a framework for engineering an adaptive, pandemic-ready health system.

### Value added

It provides an opportunity to explore the lived experience of health professionals involved in the pandemic response and to know about the challenges and gaps in the current Australian health system. Pandemic frontline healthcare workers’ perspectives will be explored in our ability to prepare the health system for future emergencies. The information will provide some insight into the engineering of a turbulence-ready health system and the digital infrastructure that will underpin it, to make the delivery of care more efficient during future crises. This study will build evidence to guide the development of future health systems adaptive to crises using digital technology.

## Data Availability

All data produced in the present study are available upon reasonable request to the authors

## Notes

### Competing Interest Statement

The authors have declared no competing interest.

### Funding Statement

This work was supported by the Australian National Health and Medical Research Council: Partnership Centre for Health System Sustainability; and Centre for Research Excellence in Digital Health
(APP1134919). The funding source did not play any role in study design, in the collection, analysis, and interpretation of data, in the writing of the report, or in the decision to submit the article for publication.

### Author Declarations

Ethics committee of Macquarie University Ethics Committee gave ethical approval for this work (Reference No: 520221118736853).

